# Range of motion at baseline predict patient reported outcome measures in frozen shoulder patients treated with injections and rehabilitation

**DOI:** 10.1101/2022.11.25.22282753

**Authors:** fabrizio brindisino, elena silvestri, Chiara Gallo, Giovanni Di Giacomo

## Abstract

Frozen shoulder is a common shoulder concern with a prevalence of 2-5 per cent in the general population that affects the shoulder joint between the ages of 40 and 60, mostly in female subjects, manifesting in progressive loss of glenohumeral movements associated with intense pain.

The pathological process consists of a fibroproliferative tissue fibrosis and an inflammation of the synovial membrane. Although the pathophysiology of this condition has been deeply studied, the mechanisms underpinning remain poorly understood.

Frozen shoulder manifests clinically as shoulder pain with progressive restricted movement, both active and passive, in the absence of trauma, along with normal radiographic scans of the glenohumeral joint.

It classically progresses through 3 overlapping stages of pain (stage 1, lasting 2-9 months), stiffness (stage 2, lasting 4-12 months) and recovery (stage 3, lasting 5-24 months); however, up to day seems that pain-predominant and stiff-predominant phases could be more usefull in treatment modality choice and managing.

The medical management has not been defined with a wide spectrum of operative and nonoperative treatments available. The most widely used treatments are local steroid and/or anesthetic injections, stretching, active and passive mobilization, physiotherapy, hydrodistension, capsular release; hoverver, he goals of the treatment are pain management, shoulder function restoration and improvement in quality of life. Based on the best available evidence it appears that the use of corticosteroid injections plus physiotherapy has been associated with better outcomes above all in terms of early benefit in ER ROM with clinical significance as long as 6 weeks of treatments.

Nevertheless, it remains unclear which parameters influence the prognosis of the pathology.

## AIM

So that, the aim of this study was to investigate whether a range of motion superior to 90 degree forward flexion in scapular plane, External Rotation (ER) at arm by side (lower than 30 per cent, between 30 and 60 per cent, 90 per cent or more than contralateral side) and abduction (lower than 30 per cent, between 30 and 60 per cent, 90 per cent or more than contralateral side) at baseline would predict best treatment outcome as measured by ROM, SPADI, NRS for Pain (night, at rest and in activity) in patients diagnosed with frozen shoulder

## HYPOTHESIS

Our hypothesis was that flexion <90°, external rotation and abduction < 30%, <60%, 90% of contralateral side could define a worst prognosis of patients outcome in frozen shoulder as measured by ROM, SPADI, NRS for Pain (night, at rest and in activity).

## POPULATION

Participants in this study were recruited by the researchers via an orthopedic surgeon. For this cohort study individuals were included when they were adults with unilateral clinically diagnosed idiopathic FS and who had not carried out any treatment prior to the baseline.

### The inclusion criteria were as follows

a. adult population ≥ 40-60 years old,
b. both male or female,
c. diagnosis of frozen shoulder contracture syndrome.
d. onset of symptoms < 3 months

### The exclusion criteria were as follows

a. patients with shoulder fractures in the last year,
b. patients with traumatic or massive rotator cuff tears in the last year,
c. patients with previous shoulder surgery procedure in the last year,
d. patients with previous shoulder dislocation in the last year,
e. patients with neoplasm,
f. patients with infections and related symptoms,
g. patients with psychiatric diagnosed disorders.

A written consent form was signed by all patients.

## BASELINE EVALUATION AND MAIN OUTCOME MEASURES

At baseline and follow-up, the following reliable and valid outcomes were performed divided into physical examination and PROMs administration:

### Physical examination

- Arm function (disability of the Arm Shoulder and Hand Questionnaire - DASH) *Disabilities of the Arm, Shoulder, and Hand Questionnaire (DASH)*: (Padua et al, 2003) The DASH is composed of 30 questions measuring physical function, social function, and symptoms in patients with musculoskeletal disorders of the upper limb. Each item is rated with a 5-points Likert scale. As the DASH Italian version (Padua et al., 2003) demonstrated a three-factors structure (Franchignoni et al., 2010), three different scores will be calculated: items 1–5, 7–11, 16–18, 20, and 21 composed the Manual Functioning subscale, items 6, 12– 15, and 19 composed the Shoulder Range of Motion subscale, and items 22–30 composed the Symptoms and Consequences subscale. The total score of each subscale was computed by adding the scores attributed to each item
- Range of Motion (ROM),

### PROMs administration

- NRS for pain at rest, night and in activity, *Pain intensity (numeric pain rating scale – NRS-pain) (Hawker et al*., *2011)* The NPRS is an 11-point numeric rating scale, to indicate the shoulder pain intensity, with 0 = no pain at all and 10 = worst imaginable pain. Participants were asked to score their average shoulder pain intensity during the present week at rest, at night and while performing activities of daily living.
- EURO-QoL 5D 5L, quality of Life scale.

## METHODS

The patients were treated with unguided intra-articular corticosteroid and anesthetic injections, painful/end-range mobilization techniques, and painful/end-range home stretching exercises.

Physical examination and PROM administration were re-administered at 3 months follow-up

The treatment consisted of an integration of technique as described in Venturin et al study of 2021 based on: 3 unguided corticosteroid/anesthetic injection throughout the rehabilitation process, performed by an orthopedist highly experienced in intra-articular shoulder injection. The first injection was administered at baseline, while the subsequent 2 were delivered at a distance of 16 weeks from the first.

The patients underwent physical therapy consisting of 6 painful/end-range mobilization techniques at the limit of each patient’s tolerance. The patients were instructed to alert the physical therapist if the excessive pain would not allow them to hold the position. These techniques were performed by the physical therapist in 30-minutes sessions, twice per week, during the first 2 weeks and once a week during the following 3 months. Patients were also taught 4 stretching ROM exercises and were instructed to perform them at home for 30 minutes, twice per day, throughout the 3 months. The patients were advised to note in a diary the exercises performed in order to ensure their execution. The choice of the treatment techniques, frequency, and type of home exercises was based on the main ROM limitations highlighted by the passive ROM examination.

The treatment supervised by the physical therapist lasted 3 months, which is the time needed to prevent any re-exacerbation of the condition.

### Study Design

Prospective cohort study

### Study Duration

Each subject’s participation will last at maximum 3 months; evaluation will be extended for 6 months.

### Study Phases Screening

Screening for eligibility: the patients will be screened for inclusion and exclusion criteria and will be informed about the study procedures and aims. All patients will sign a written informed consent to the study.

### Study Treatment

The treatment will be the best choice up today highlighted in literature (corticosteroid infiltrations, mobilization, stretching, exercises) and no additional intervention will be administered.

### Follow-Up

Intermediate follow up will be as shown in Table 1

**Table 1.**
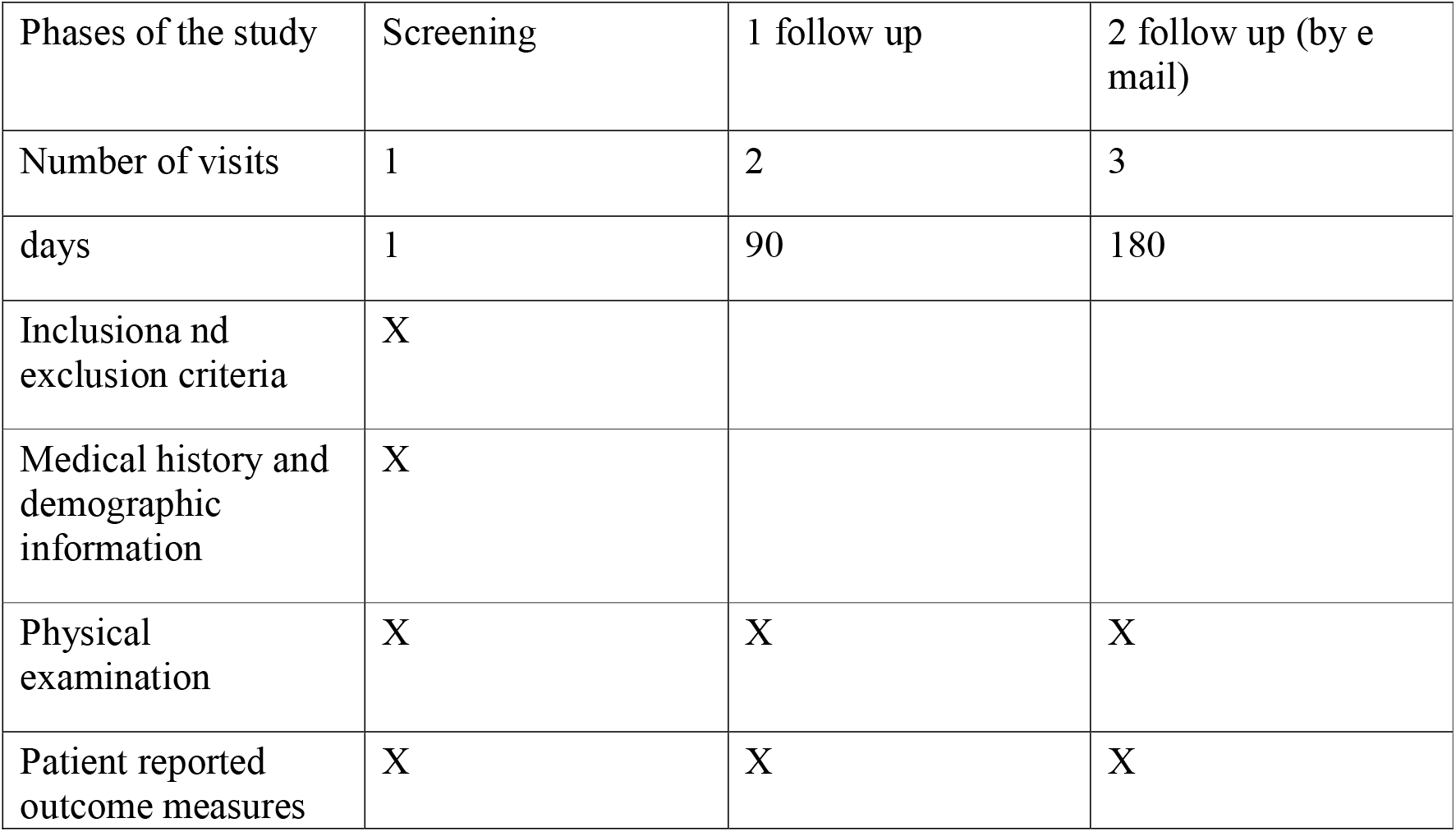
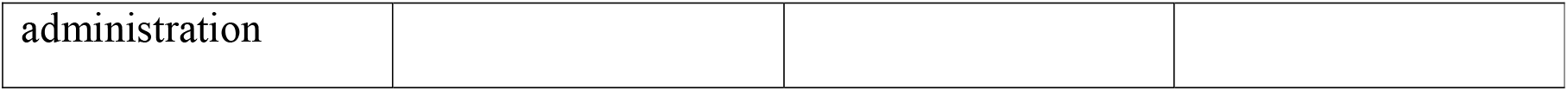
study’s synopsis

All statistical analyzes will be performed with SPSS software

## Data Availability

All data produced in the present study are available upon reasonable request to the authors

